# Local prevalence of ceftriaxone resistance informs optimal deployment of new gonorrhea treatments

**DOI:** 10.64898/2026.04.23.26351610

**Authors:** Kirstin I. Oliveira Roster, Minttu M. Rönn, Eva R. Gorenburg, Dennis K. Partl, Nanina Anderegg, Pia Abel zur Wiesch, Carmen Au, Roger D. Kouyos, Fernando P. Martinez, Nicola Low, Yonatan H. Grad

## Abstract

Numerous factors may influence the optimal rollout of new gonococcal antibiotics. We compared eight rollout strategies using a gonorrhea transmission model and ranked strategies by the number of gonococcal infections and clinically useful antibiotic lifespan. Rankings were most sensitive to the starting ceftriaxone resistance prevalence and screening frequency.

## Introduction

*Neisseria gonorrhoeae* has developed resistance to all antibiotics recommended for first-line treatment. The prevalence of resistance to ceftriaxone, the mainstay of therapy, is rising globally (1). Controlling gonorrhea is challenging, especially in low- and middle-income countries (LMICs), with men who have sex with men (MSM) a key population disproportionately affected by gonorrhea. In much of the world, data on the prevalence of resistance are sparse (2,3), and a lack of diagnostics means sexually transmitted infections (STIs) are managed syndromically (4).

Increasing access to novel therapeutics and diagnostics is paramount to improving global health and preventing the spread of resistance (5–7). Two new antibiotics, gepotidacin (8) and zoliflodacin (9), received approval from the US Food and Drug Administration in December 2025 for treatment of gonorrhea; the rollout of zoliflodacin includes a focus on LMICs (10). The approval of Nuzolvence® (zoliflodacin) and its recommended label use (11), although broad reflects uncertainties about potential reproductive effects observed in animal studies, which suggest that early rollout may initially focus on MSM not interested in reproduction. Ongoing studies are generating data that will provide additional confidence in the use of the antibiotic among populations already included in the broad label. Until such data are available, the optimal strategy for rollout in MSM, and how the strategy should be shaped by local circumstances, is unclear.

Here, we aimed to address these gaps by comparing the performance of rollout strategies with varying ceftriaxone resistance, sexual behaviors, and screening in a model of gonorrhea transmission in MSM.

## Methods

We adapted a gonorrhea transmission model in MSM, initially developed for the US population (12), and compared several strategies for rolling out a new antibiotic in their ability to lower gonorrhea incidence and extend the clinically useful lifespan of antibiotics. The mathematical model incorporates sexual partner change rates, risk heterogeneity, and sexual mixing, and accounts for symptomatic and asymptomatic infections with four strains of *N. gonorrhoeae*: sensitive to treatment with ceftriaxone and the new antibiotic (termed antibiotic A), ceftriaxone-resistant, resistant to antibiotic A, and resistant to both antibiotics.

We simulated eight strategies for the introduction of antibiotic A: 1) all diagnosed infections treated with ceftriaxone and antibiotic A (Combination therapy); 2) 50% of cases treated with antibiotic A, the rest with ceftriaxone (Random 50%); 3) 20% of cases treated with antibiotic A, the rest with ceftriaxone (Random 20%); 4) antibiotic A introduced gradually over 8 years, switching from first-line ceftriaxone to antibiotic A (Gradual 100%); 5) antibiotic A introduced gradually over 8 years, with 50% of cases treated with antibiotic A and 50% with ceftriaxone by the end of roll-out (Gradual 50%); 6) antibiotic A reserved as second line treatment until ceftriaxone resistance prevalence ≥5% of all infections, then antibiotic A is introduced over 1 year as first-line therapy (Reserve, quick 100%); 7) antibiotic A reserved until ceftriaxone resistance prevalence ≥5%, then change over 8 years from ceftriaxone to antibiotic A (Reserve, gradual 100%), 8) antibiotic A reserved until ceftriaxone resistance prevalence ≥5%, then change over 8 years from ceftriaxone to treatment of 50% of cases with antibiotic A and 50% with ceftriaxone (Reserve – gradual 50%).

While the baseline scenario reflects a model calibrated to gonorrhea epidemiology among US MSM, we varied key parameters to explore epidemiological and resource contexts. We conducted broad parameter sweeps over the starting prevalence of ceftriaxone resistance, gonorrhea screening frequency if asymptomatic, proportion of infections with symptoms (interpreted as the likelihood of seeking care if symptomatic), partner change rates, levels of assortativity (like-with-like mixing in relation to sexual activity), and number of sexual activity groups (population variation in sexual activity; **Supplemental Material**). We conducted sensitivity analyses, varying the emergence of resistance and fitness cost of resistance for each antibiotic.

For each strategy, we estimated 1) the total number of gonococcal infections over the model simulation period (60 years) and 2) the clinically useful lifespan of each antibiotic, defined as the time until resistance prevalence reaches 5% of all infections. We ranked strategies by performance for each outcome and then took the average of these ranks to obtain a composite ranking.

## Results

The cumulative incidence and antibiotic lifespan were sensitive to starting prevalence of ceftriaxone resistance, asymptomatic screening frequency and proportion of symptomatic infections (**Figure 1**). In settings with low ceftriaxone resistance, combination therapy was the most favorable strategy. With higher levels of resistance, the effective lifespan of the antibiotics was shortened, which lowered the overall composite ranking. In these high-resistance settings, strategies that allocated treatment equally (50% each) between ceftriaxone and antibiotic A were more favorable, because antibiotic A had a longer useful lifespan under these conditions (**Figure 1A**). The asymptomatic screening frequency influenced the optimal rollout strategy. When ≤40% of the modeled population was screened per year, combination therapy (strategy 1) was optimal. This strategy’s composite ranking dropped slightly with higher screening frequency, driven by a reduced lifespan of the antibiotics. When the entire modeled population was screened annually, treating 20% (strategy 3) of diagnosed infected individuals with the new antibiotic was the optimal strategy (**Figure 1B**).

**Figure 1.**
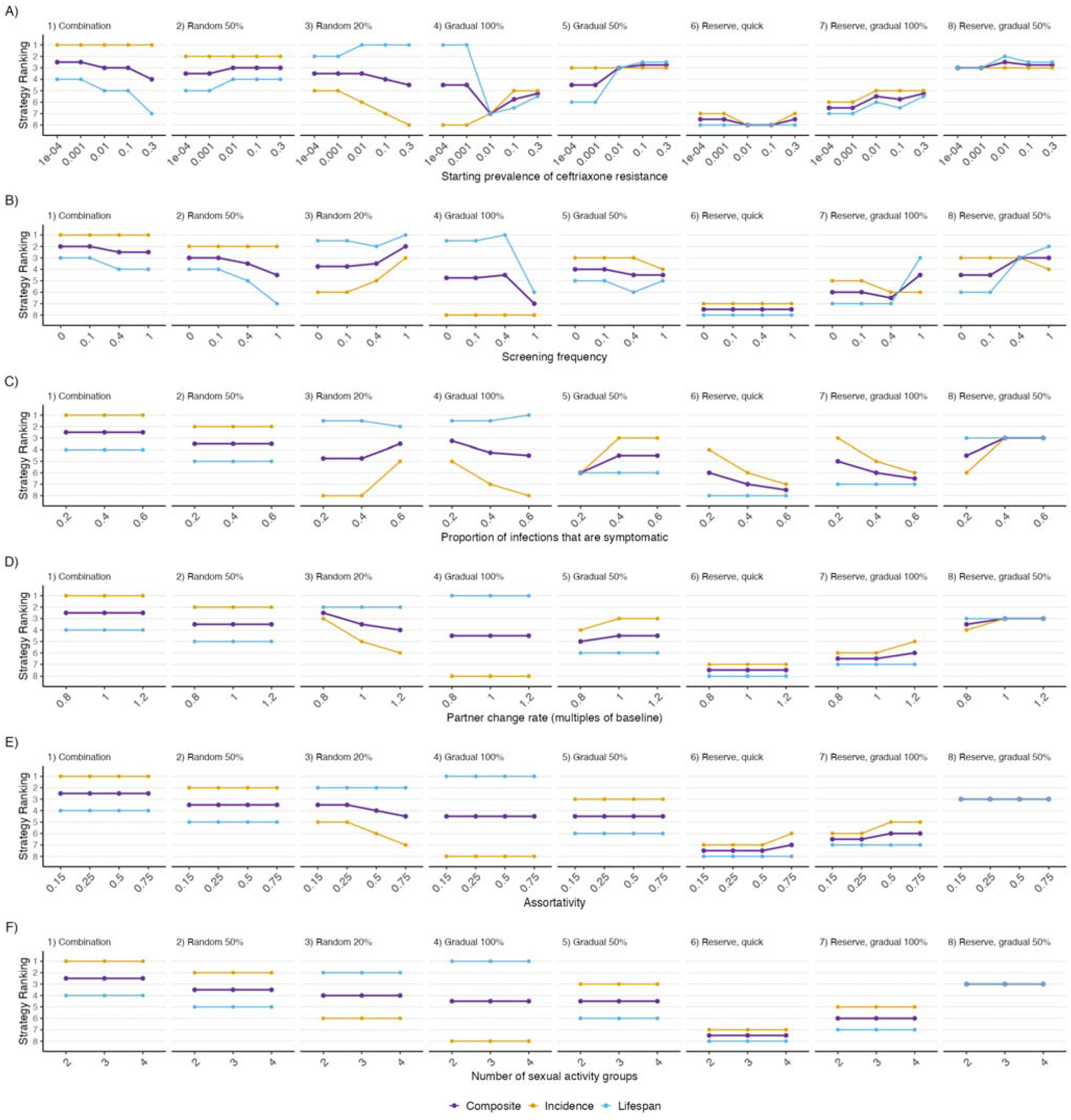
Ranking of antibiotic rollout strategies for varying parameter values. Strategies are ranked best (top) to worst (bottom) on the y axis with each strategy presented in a separate panel. Lines represent ranking by cumulative incidence (yellow), and by antibiotic lifespan (light blue), and their composite (average) ranking (dark purple). The parameters examined are on the x-axis for A) the starting prevalence of ceftriaxone resistance (proportion of infections that are ceftriaxone resistant), B) the asymptomatic screening frequency, C) the symptomatic proportion of gonococcal infections, D) multiples of partner change rates, E) measure of assortative mixing among one’s own sexual activity group, and F) the number of sexual activity population subgroups stratified by partner change rates.

Similarly, a high proportion (60%) of symptomatic infection altered the ranking. Other parameters, including aggregate characteristics of sexual behavior, had less influence on strategy rankings (**Figure 1**).

In sensitivity analyses, we varied the likelihood of emergence of resistance and the fitness cost of resistance for each antibiotic (**Supplementary Figures 1-3**). As in our main analysis, the starting prevalence of ceftriaxone resistance altered strategy rankings, but the likelihood that resistance emerged upon treatment had minimal impact (**Supplementary Figure 1**). Across resistance emergence probabilities, the optimal strategy based on the composite ranking was either combination therapy (strategy 1) or the fast reserve strategy (strategy 6) when the starting prevalence of ceftriaxone resistance was low, and random allocation (50%; strategy 2) or gradual introduction (50% of the population; strategy 5) when ceftriaxone resistance was high.

The relative fitness of ceftriaxone and antibiotic A resistant strains had a larger impact on the optimal strategy. If relative fitness of both resistant strains was much lower than in the main analysis, then gradual introduction (50% of the population; strategy 8) of antibiotic A was the optimal strategy based on the composite ranking with a low starting prevalence of ceftriaxone resistance, and random allocation (20%; strategy 3) was the optimal strategy when starting with a high prevalence of ceftriaxone resistance. When the relative fitness of either resistant strain was similar to that in the main analysis, combination therapy (strategy 1) was most often the optimal strategy (**Supplementary Figure 2**).

## Discussion

This analysis used simulations of gonorrhea transmission in MSM to identify parameters that will be critical inputs into the decision-making process for new antibiotic deployment. We found that the initial prevalence of ceftriaxone resistance and the frequency of screening for asymptomatic infection had the greatest influence on the outcomes of the antibiotic rollout strategies, while the proportion of symptomatic infections was influential at higher end of the parameter range. These parameters influence overall rates of antibiotic use, with more use of the novel antibiotic increasing opportunities for *de novo* emergence of resistance. Our results suggest that introduction of new antibiotics should take the local context into account, given that the prevalence of ceftriaxone resistance is estimated at around 30% of infections in parts of East Asia (13) but only sporadic cases have been reported in other regions (1).

This study is based on a US MSM model of *N. gonorrhoeae* transmission. The parameter ranges explored in the analysis covered a broad range of epidemiological scenarios for natural history, prevalence of ceftriaxone resistance, and sexual behavior, reflecting conditions that may occur in varying settings, including LMICs. Calibration to country-specific data would enable a tailored analysis. The optimal strategy is less influenced by sexual behavior, which suggests that the results are generalizable across MSM populations with different levels of sexual activity and sexual network structures.

While we focused on MSM (11), future analyses and models should consider strategies in heterosexual populations, once ongoing studies inform broader use, and more complex strategies that account for heterogeneity in healthcare access and health system realities. This includes evaluating the influence of other parameters, such as partner concurrency, interactions between heterosexual and MSM networks, and the duration of asymptomatic infections.

New gonorrhea treatments represent an important opportunity to reduce the global burden of gonorrhea and limit AMR with carefully designed and context-specific treatment guidelines. Our findings using a model of gonorrhea transmission in MSM highlight that prevalence of ceftriaxone resistance and asymptomatic screening frequency are key factors in selecting the drug rollout strategy to minimize resistance and burden of disease.

## Data Availability

All data produced in the present study are available upon reasonable request to the authors

## Acknowledgements

This study was funded by the Global Antibiotic Research and Development Partnership (to YHG). KIOR and YHG conceived the study. YHG obtained the funding. KIOR developed the model, led the analysis, and drafted the manuscript. MMR supported the analysis and edited the manuscript. ERG, DKP, NA, PAzW, CA, RDK, FPM, NL, and YHG provided feedback on the analysis plan, discussed the results, and contributed to the final manuscript. CA and FPM are employees of the Global Antibiotic Research and Development Partnership, which funded this study. YHG has consulted for the Analysis Group and GSK on topics not related to this study and is on the scientific advisory board of Kanso Diagnostics. No other conflicts of interests were declared.

