## Supplemental Material for "Local prevalence of ceftriaxone resistance informs optimal deployment of new gonorrhea treatments"

### Supplementary Materials

#### I. Supplementary Results

**Figure S1. Sensitivity analysis of strategy rankings for varying combinations of resistance emergence probabilities against antibiotic A and ceftriaxone.**

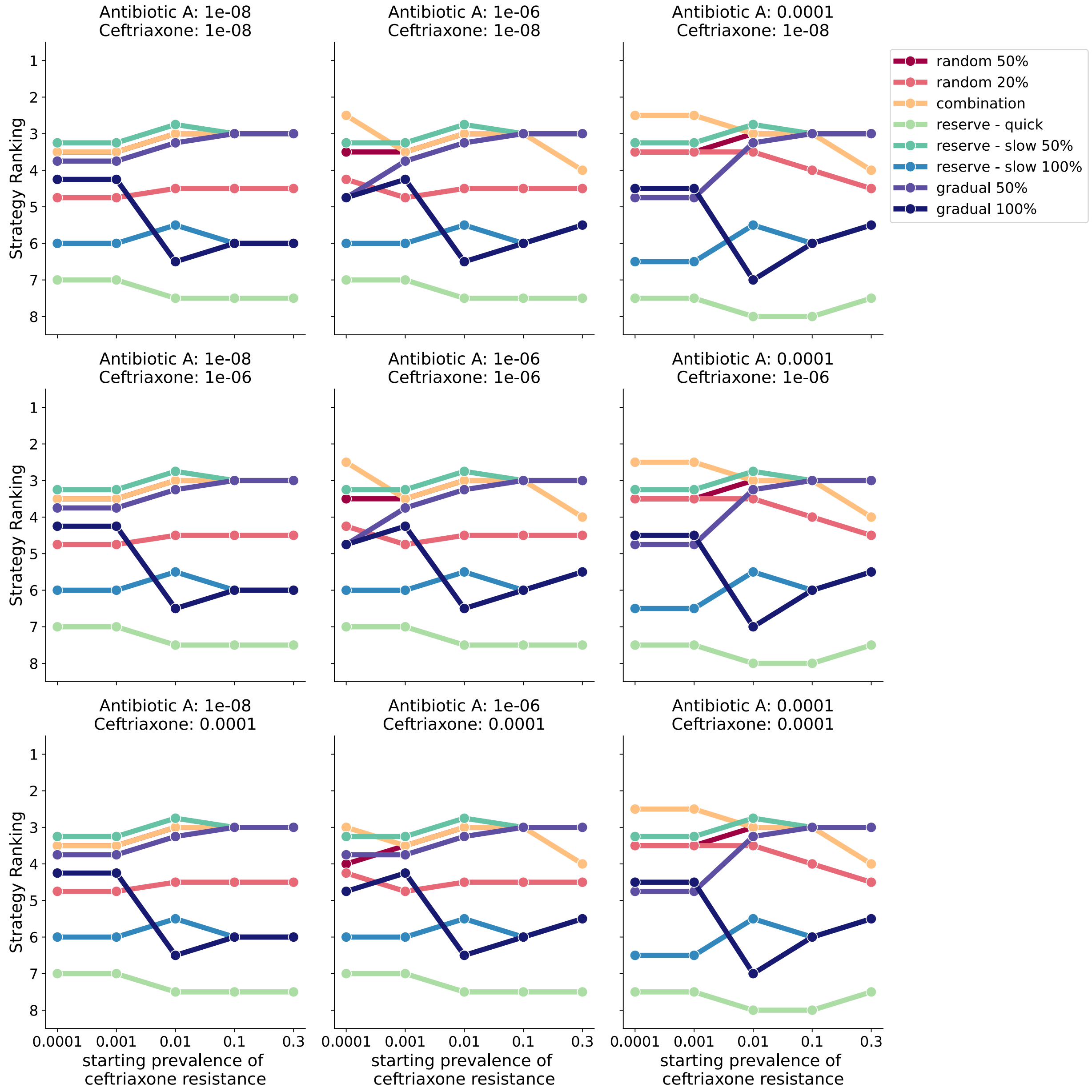

**Figure S2. Sensitivity analysis for varying values of relative fitness of ceftriaxone-resistant and antibiotic A-resistant strains.**

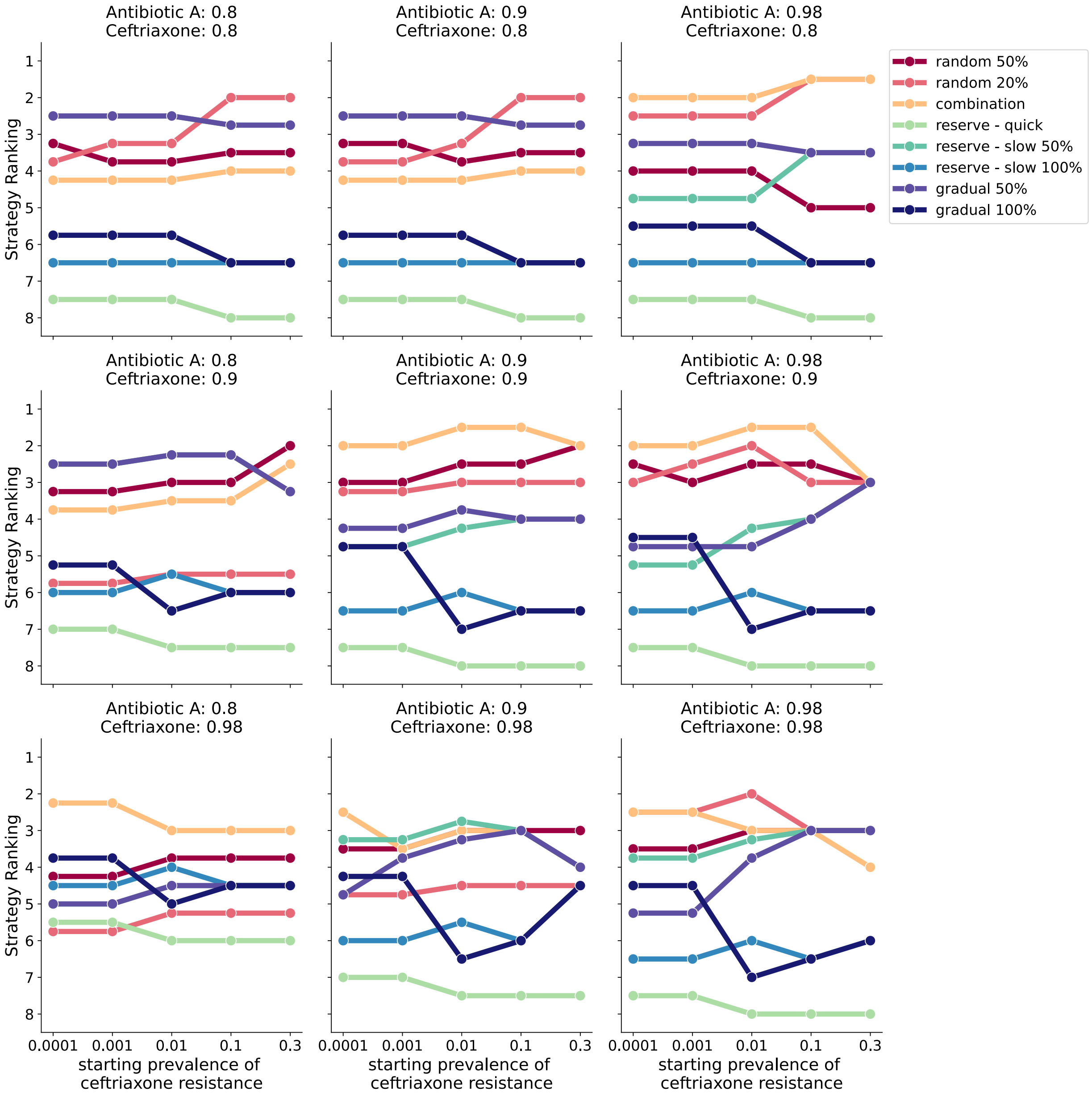

### Figure S3. Ranking antibiotic rollout strategies for individual outcome measures

#### A) Total infections

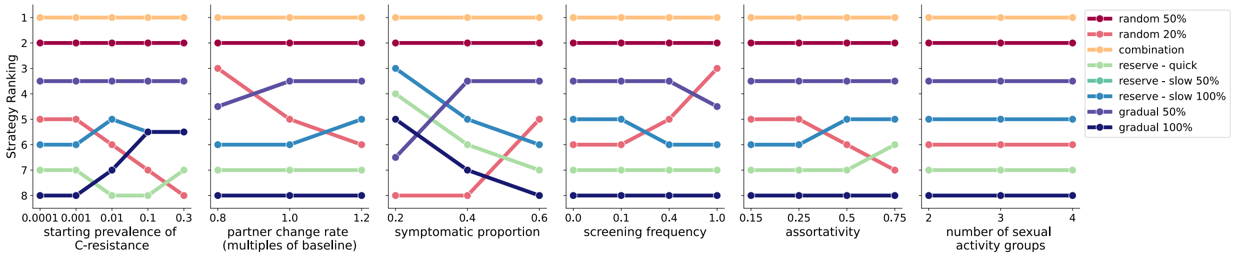


#### B) Clinically useful antibiotic lifespan

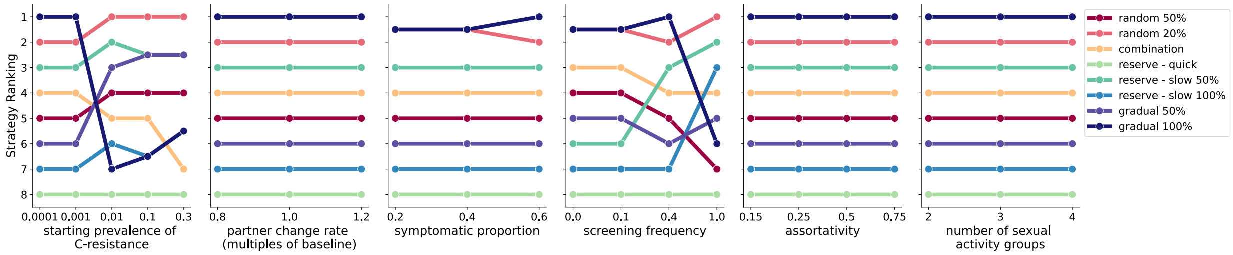

#### II. Supplementary Methods

Compartmental Model Structure

We adapted a published and validated compartmental model to simulate the transmission of gonorrhea among 1 million men who have sex with men (MSM) (1). The model accounts for both symptomatic (Y) and asymptomatic (Z) infections with four strains of *N. gonorrhoeae*: strains that are sensitive to treatment with ceftriaxone and the new antibiotic (antibiotic A) ($Y_0$, $Z_0$), ceftriaxone-resistant ($Y_c$, $Z_c$), antibiotic A-resistant ($Y_A$, $Z_A$), and resistant to both antibiotics ($Y_{cA}$, $Z_{cA}$).

Initially, a small proportion of infections was ceftriaxone-resistant, and the rest was sensitive to treatment with both drugs. Resistance emerged upon treatment and was spread through transmission and selection for resistant strains, which evade treatment with either ceftriaxone, antibiotic A, or both.

Recovery occurred either naturally, through successful treatment, or through retreatment if the first treatment was unsuccessful. We assumed that a last-line antibiotic is used for retreatment and successfully cleared all infections with any strain.

The population was further stratified by partner change rates into three sexual activity groups.

The baseline parameters guiding these transitions in the baseline analysis were taken from Reichert et al. (2023) (1) and are replicated in **Table S1**.

All code required to reproduce the analyses is available at https://github.com/KRoster/lmic_gc.

**Table S1. Baseline Parameters.**

| **Parameter** | **Value** |
|---|---|
| Gonorrhea prevalence at start of simulation (and calibration target) | 3% |
| Population size | 1 million |
| Relative size of sexual activity groups:<br>- Low<br>- Medium<br>- High | <br>- 30%<br>- 60%<br>- 10% |
| Number of partners per year per sexual activity group<br>- Low<br>- Medium<br>- High | <br>- 1*1.22<br>- 5*1.22<br>- 20*1.22 |
| Mixing parameter among sexual activity groups (assortativity) | 0.24 |
| Proportion of infections that are resistant to ceftriaxone at start of simulation | 0.0001 |
| Proportion of infections that are resistant to antibiotic A at start of simulation | 0 |
| Model entry and exit rate (per year) | 1/20 |
| Proportion of incident infections that are symptomatic | 0.6 |
| Transmission probability per partnership | 0.46 |
| Time until natural recovery, in days | 168.6 |
| Time until treatment of symptomatic infections, in days | 11.3 |

| | |
|---|---|
| Time until retreatment if initial treatment of symptomatic infection fails, in days | 11.3*3 |
| Asymptomatic screening rate (per year) | 0.4 |
| Probability of emergence of resistance upon treatment<br>- Ceftriaxone resistance<br>- Antibiotic A resistance<br>- Dual resistance | <br>- $10^{-8}$<br>- $10^{-4}$<br>- $10^{-8} * 10^{-4}$ |
| Relative fitness of resistant strains, compared to sensitive:<br>- Ceftriaxone resistance<br>- Antibiotic A resistance<br>- Dual resistance | <br>- 0.98<br>- 0.95<br>- 0.98 * 0.95 |

<u>Antibiotic A rollout strategies</u>

We simulated eight strategies for the introduction of antibiotic A:

- ***Combination therapy***: all infections are treated with both ceftriaxone and antibiotic A
- ***Random 50%*****:** 50% of cases are treated with antibiotic A, the rest with ceftriaxone
- ***Random 20%:*** 20% of cases are treated with antibiotic A, the rest with ceftriaxone
- ***Gradual 100%:*** antibiotic A is gradually introduced, such that over a period of 8 years, treatment switches from treatment of all cases with ceftriaxone to treatment of all cases with antibiotic A
- ***Gradual 50%:*** Antibiotic A is gradually introduced, such that over a period of 8 years, treatment switches from treatment of all cases with ceftriaxone to treatment of 50% of cases with antibiotic A and 50% with ceftriaxone
- ***Reserve – quick:*** Antibiotic A is held in reserve until the prevalence of ceftriaxone resistance reaches at least 5% of all infections, at which point antibiotic A is introduced quickly, such that over a period of 1 year, treatment switches from ceftriaxone to antibiotic A
- ***Reserve – gradual 100%:*** Antibiotic A is held in reserve until the prevalence of ceftriaxone resistance reaches at least 5% of all infections, at which point antibiotic A is introduced gradually, such that over a period of 8 years, treatment switches from ceftriaxone to antibiotic A
- ***Reserve – gradual 50%:*** Antibiotic A is held in reserve until the prevalence of ceftriaxone resistance reaches at least 5% of all infections, at which point antibiotic A is introduced gradually, such that over a period of 8 years, treatment switches from ceftriaxone for all cases to antibiotic A for 50% of cases and ceftriaxone for the remaining 50%

<u>Simulation of country context</u>

We simulated variation in six parameters representing epidemiological, behavioral, and health systems differences across countries. Specifically, we varied:
(a) the starting prevalence of ceftriaxone resistance, considering settings where 0.01%, 0.1%, 1%, 10%, and 30% of cases are resistant to ceftriaxone at the start of the simulation
(b) partner change rates, where the baseline partner change rates in the three sexual activity groups (**Table S1**) are scaled up or down by factors 0.8 (lower activity), 1.0 (baseline levels), and 1.2 (higher activity),
(c) the proportion of gonococcal infections that are symptomatic, considering scenarios where 20%, 40% or 60% of all infections lead to symptoms,

(d) the asymptomatic screening frequency, considering settings where there is no asymptomatic screening at all (0 screenings per year) or where each person in the simulation is screened 0.1, 0.4, or 1.0 times per year,
(e) the assortativity of mixing among sexual activity groups, defined as the propensity of individuals to mix with others in the same sexual activity group, with values 0.15, 0.25, 0.5, and 0.75, and
(f) the number of sexual activity population subgroups stratified by partner change rates, while maintaining the same population-aggregate level of sexual activity, but dividing partnerships among 2, 3, or 4 groups with adjusted relative sizes and partner change rates (**Table S2, Table S3**).

**Table S2. Sizes of sexual activity groups in different population heterogeneity scenarios.**

| Scenario | Very high activity | High activity | Medium activity | Low activity |
|---|---|---|---|---|
| 2 groups | | 10% | | 90% |
| 3 groups | | 10% | 60% | 30% |
| 4 groups | 2% | 8% | 60% | 30% |

**Table S3. Relative partner change rates in different population heterogeneity scenarios.**

| Scenario | Very high activity | High activity | Medium activity | Low activity |
|---|---|---|---|---|
| 2 groups | | $k_h$ | | $1/3^* k_l + 2/3 * k_m$ |
| 3 groups | | $k_h$ | $k_m$ | $k_l$ |
| 4 groups | $1.4^* k_h$ | $0.9^* k_h$ | $k_m$ | $k_l$ |

<u>Statistical analysis</u>

We ran each simulation for 60 years and calculated (i) the cumulative number of gonococcal infections that occurred over this time period and (ii) the antibiotic lifespan, defined as the time in years until prevalence of resistance to both antibiotics reaches at least 5% of all infections.
